# Covariate clustering: women with breast cancer in southwestern Paraná, Brazil

**DOI:** 10.1101/2021.10.04.21264299

**Authors:** Neyva M. L. Romeiro, Carolina Panis, Mara C.T. dos Santos, Daniel Rech, Paulo L. Natti, Eliandro R. Cirilo

**Affiliations:** Mathematics Department, Universidade Estadual de Londrina – UEL, Londrina, PR, Brazil; Laboratory of tumor biology, Universidade Estadual do Oeste do Paraná - UNIOESTE, Francisco Beltrão, Pr, Brazil; Master’s Student of the Graduate Program in Applied and Computational Mathematics – PGMAC, Universidade Estadual de Londrina – UEL, Londrina, PR, Brazil

**Keywords:** Breast Cancer, Paraná-Brazil, Clusters, Body Mass Index (BMI), TNM Staging, Menopause

## Abstract

Due to the high incidence and aggressiveness of breast cancer, the understanding of specific factors associated with the profile of the disease is necessary. In this context, the aim of the study was to analyze data from 155 patients with breast cancer, attended at a reference hospital for Oncology of the Unified Health System (SUS), in the period 2015-2020, in the southwest region of Paraná, Brazil. Using multivariate statistical analysis, sample data were divided into three clusters. The heterogeneity between clusters was obtained by Ward’s method. The clinical and pathological variables obtained from the patients’ medical records were: presence of intratumoral emboli, presence of lymph nodes, menopausal status, molecular subtype of breast cancer, histological grade, TNM staging of the disease, tumor size (cm), age at diagnosis (years), weight (kg), height (m2) and body mass index (BMI) (kg/m^2^). From the data of the total sample, it is observed that 70% of the patients were in menopause at diagnosis, 31.5% had tumors containing emboli, and 41% had positive lymph nodes. The prevalence of Luminal subtype B tumors, intermediate histological grade, and TNM staging II was verified. Furthermore, the prevalence of the disease was higher in women aged over 50 years, representing 66% of cases. The BMI of the patients ranged from 17.63 kg/m^2^ to 51.26 kg/m^2^, with 26.45% of the patients with a BMI below 25 kg/m^2^, 40.65% with a BMI between 25 kg/m^2^ and 30 kg/m^2^ and 32.9% with BMI above 30 kg/m^2^. Cluster analysis, using the spatial distribution of patients, showed that the region of Vale do Iguaçu was the region with the worst averages for clinical-pathological variables, while the region of Vale do Marrecas had the highest number of breast cancer cases.

## INTRODUCTION

Breast cancer is the most common malignant neoplasm in women [1]. Age, over 50 years old, is the most important risk factor [2]. Other determining factors for the development of the disease are genetic, hereditary, late menopause, obesity, sedentary lifestyle and frequent exposure to ionizing radiation [3,4]. Such factors are mainly responsible for the clinicopathological differences found in the literature on breast cancer [5-11].

Specific studies involving the Brazilian population point to the occurrence of classic risk factors, such as aging and menopausal status [12,13]. Other studies show more complex associations, also observed around the world, such as the development of tumors with a worse prognosis, such as triple negative, in obese and overweight women [11,14]. Factors such as social vulnerability [15] and a history of psychological stress [16] have also been reported as possible risks associated with the presence of breast cancer in women living in southern Brazil. However, studies referring to regional risk factors are rare and not very conclusive.

In this context, it is intended to categorize, through statistical analysis, possible risk factors for breast cancer, targeting patients in the southwest region of Paraná, Brazil. It is known that mathematical analysis can be a powerful tool to assess patient data, providing reliable associations between variables that often cannot be understood in isolation. Considering, further, that physicians may not be familiar with statistical analysis, such interdisciplinary studies become essential.

Data from breast cancer patients can be analyzed using various mathematical tools. We highlight the multivariate analysis that studies the correlation of two or more variables with different information [17-19]. Data clustering, one of the fields of multivariate statistical analysis, divides sample data into groups with correlated elements. The analysis of these groups can provide relevant information about part of the total sample. In this way, clustering performs a more specific descriptive analysis of the groups within the sample.

In the literature, it is observed that many studies perform statistical analysis considering correlations of few variables [5,8-11,20]. In this line of study, to categorize possible risk factors identified in women diagnosed with breast cancer in the 8th Health Regional of the State of Paraná, an exploratory data study is presented considering 11 clinical and pathological variables. For this study, the Euclidean distance between the variables is calculated, which makes it possible to determine the sequence of data grouping. Then we apply Ward’s hierarchical method that identifies the clusters. The Means Test will be used to verify significant differences between the means of the variables in the clusters. Finally, Spearman’s rank correlation analysis measures, in the general data of the sample and in the clusters, the “strength” that one variable applies to the others.

## METHODS

### Sample

The sequential data used contain information from biopsy samples taken serially from women who had lesions suggestive of breast cancer, visualized by imaging tests (mammography, ultrasound, or MRI) and physical examinations, in the period from May 2015 to March 2020. Data confidentiality was maintained in accordance with clinical research guidelines. The study was approved by the Institutional Ethics Board of Western Paraná State University, through Plataforma Brasil, under the number CAAE 35524814.4.0000.0107, where CAAE stands for Certificate of Presentation of Ethical Appreciation. The study included 155 patients with a confirmed diagnosis of breast cancer through biopsy. These patients from the 8th Health Regional of the State of Paraná, which covers 25 municipalities, were treated at the Francisco Beltrão Cancer Hospital, in the city of Francisco Beltrão, Paraná, Brazil. The 25 municipalities were divided into three regions, with 35.5% of the patients living in the Fronteira region, 20% in the Vale do Iguaçu region, and 44.5% in the Vale do Marrecas region, as illustrated in Figure 1 (a)-(c), respectively. Medical records were consulted to obtain data.

**Figure 1.**
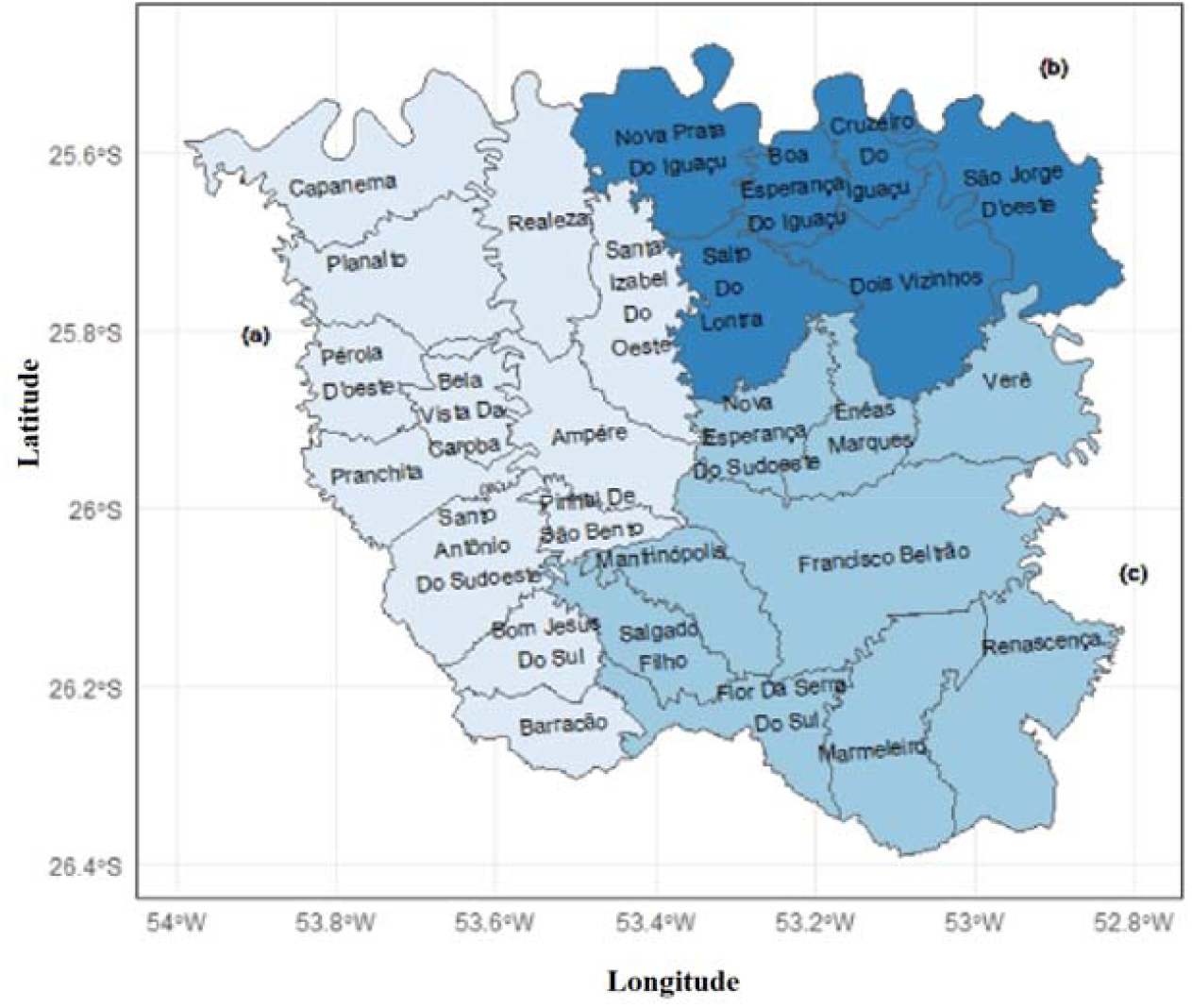
Municipalities of the 8th Health Regional of the State of Paraná, Brazil: (a) Fronteira Region, (b) Vale do Iguaçu Region, and (c) Vale do Marrecas Region. Source: Adapted from http://paginapessoal.utfpr.edu.br/fernandoramme/mapas/sudoeste/pdri2013sudoeste.png

All patients signed consent and each protocol followed the principles of medical research involving humans described in the Declaration of Helsinki.

### Variables

In this study, variables with different characteristics and applications are considered. The histopathological variable was used to verify which of the patients in the sample were confirmed with breast cancer. The region variable (location) was used to verify the spatial distribution of breast cancer cases by the municipalities of the 8th Health Regional of the State of Paraná, Figure 1. For the patient variable, we chose to label this variable, not by the name of each patient, but by numbers, which guarantees any relationship with an individual.

Furthermore, 11 clinicopathological variables are used to describe particular characteristics of the disease, such as: the presence of intratumoral emboli, the presence of lymph nodes, the menopausal status, the molecular subtype of breast cancer, the histological grade, the TNM staging of the disease, tumor size (cm), age at diagnosis (years), weight (kg), height (m) and body mass index (BMI) in (kg/m^2^).

Histopathological evaluation is essential for the diagnosis of neoplasia. In this context, for histological grade variable, the following criteria were adopted:

1. well differentiated;
2. moderately differentiated;
3. little differentiated.

For molecular subtype variable, four categories were adopted according to the following criteria:

1. Luminal subtype A: tumor that presents positivity above 1% for the expression of estrogen and/or progesterone receptors, with negative HER2 protein and ki67 protein below 14%;
2. Amplified HER2 subtype: tumor that is negative for the expression of estrogen (ER) and/or progesterone (PR) receptors and with a HER2 score greater than 2+; or even, with a HER2 score equal to 2+ and with amplification confirmed by the FISH technique associated with any ki67 value;
3. Luminal subtype B: tumor that presents positivity above 1% for the expression of estrogen and/or progesterone receptors, with negative HER2 and ki67 above 14%;
4. Triple negative subtype: tumor that presents negativity for the expression of ER, PR and HER2, regardless of the ki67 value.

The TMN staging variable was classified in relation to the stages of the disease as described by *American Joint Commitee on Cancer, Breast Cancer Staging System* [21].

The age at diagnosis variable was categorized below or above 50 years. The variable body mass index is categorized as normal BMI when below 25 kg/m^2^, overweight with a BMI between 25 and 30 kg/m^2^, or obesity with a BMI greater than 30 kg/m^2^.

For a better understanding of the work, the variables are classified as:

**Binary variables:** They describe the presence of angiolymphatic emboli, the presence of lymph node invasion, and menopausal status. These variables assume the values 0 or 1, which represents no or yes, respectively;

**Categorical variables:** They categorize possible risk factors by assessing molecular subtype, histological grade, and TNM staging. These variables aim to determine the characteristics of the tumor, the type of cancer, and even the severity of the disease;

**Quantitative variables:** These describe the tumor size, age at diagnosis, weight, height and body mass index of the patient.

### Statistical methods

Cluster analysis, or data clustering, analyzes characteristics that differentiate the data from a sample, dividing it into groups [17,19,22].

There are several methodologies that apply the cluster concept, including the Ward method. In this article, cluster analysis is performed by calculating the Euclidean distance between the set of 11 clinicopathological variables.

Initially, to determine which data are more homogeneous with each other, the Euclidean distance method is used. Next, Ward’s hierarchical agglomerative method is used to generate the heterogeneous groups among themselves. The result of the analysis, presented in the form of a dendrogram, helps to identify the division of groups, thus generating clusters.

Once the clusters were obtained, the calculation of Spearman’s linear correlation between the variables allowed us to understand the influence that one variable exerts over another, enabling the identification of possible risk factors associated with the groups [23]. The Spearman’s correlation coefficient varies between -1 (great disagreement between the groups of each pair of variables) and +1 (great agreement between the groups of each pair of variables). To determine the significance of the correlations, the p-value is calculated.

To extract characteristics that distinguish the data from different clusters, in addition to calculating the correlation, the Test of Means is used. This procedure allows us to calculate, for each of the cluster variables, those that present different significant means and those that are just sample variations.

## RESULTS AND DISCUSSION

From Table 1, approximately 31% of the patients had tumors containing angiolymphatic emboli. It is also noted that the presence of positive lymph nodes was observed in 41% of patients, and that 70% are classified as menopausal women at diagnosis.

**Table 1.** Information on the clinical-pathological variables of the sample

|  | Min Value | 1st Quad | Median | Mean | 3rd Quad | Max Value |
| --- | --- | --- | --- | --- | --- | --- |
| **Binary variables** |  |  |  |  |  |  |
| Presence of intratumoral emboli | 0 | 0 | 0 | 0,31 | 1 | 1 |
| Presence of lymph node invasion | 0 | 0 | 0 | 0,41 | 1 | 1 |
| Menopausal status | 0 | 0 | 1 | 0,70 | 1 | 1 |
| **Categorical variables** |  |  |  |  |  |  |
| Molecular subtype * | 1 | 1 | 3 | 2,43 | 3 | 4 |
| Grade * | 1 | 1 | 2 | 1,93 | 2 | 3 |
| TNM staging * | 0 | 2 | 2 | 2,06 | 2 | 4 |
| **Quantitative variables** |  |  |  |  |  |  |
| Tumor size (cm) | 0,9 | 2 | 2,5 | 3,18 | 4 | 15 |
| Age (years) | 31 | 45 | 58 | 56,6 | 66 | 96 |
| Weight (kg) | 39,3 | 63 | 71,4 | 72,5 | 82 | 120 |
| Height (m) | 1,42 | 1,56 | 1,60 | 1,60 | 1,65 | 1,80 |
| BMI (kg/m<sup>2</sup>) | 17,6 | 24,8 | 27,6 | 28,3 | 31,2 | 51,3 |

\* Molecular subtype was classified into 4 categories. Histological grade categorized into 3 criteria. TNM staging refers to stages 0, I, II, III and IV, respectively.

On average, the patients in this study have a higher frequency of tumors of the Luminal B molecular subtype, intermediate histological grade, moderately differentiated, and a median TNM stage II, with variations between 0 and IV.

There was also an important dispersion of tumor size, ranging from 0.9 cm to 15 cm. The average age of patients is 56.6 years, and the prevalence of the disease was higher in women aged over 45 years, representing 75% of cases. The average weight, when diagnosed, was close to 72.5 kg, but one of the patients weighed 120 kg. Furthermore, only 25% of patients had a BMI of less than 24.8 kg/m^2^.

### Clinicopathological correlations and identification of possible associated risk factors

#### General data of the sample

Checking the influence that one variable exerts on the other allows a better understanding of the data from a sample, which makes it possible to identify possible risk factors. Thus, Spearman’s linear correlation is used to estimate the correlation between each pair of variables, evaluating possible connections between the presence of intratumoral emboli, lymph node invasion, menopausal status at diagnosis, molecular cancer subtype, histological grade, TNM staging, tumor size, age, weight, height and BMI. These results are shown in Table 2, where the difference between the groups with p-value < 1% was considered significant.

**Table 2.** Spearman’s correlations for the 155 patients diagnosed with breast cancer.

| Variables | Presence of lymph node invasion | TNM staging | Age | Height | BMI |
| --- | --- | --- | --- | --- | --- |
| Presence of intratumoral emboli | 0,46* | 0,36* |  |  |  |
| Presence of lymph node invasion |  | 0,6* |  |  |  |
| Menopausal status |  |  | 0,7* |  |  |
| Weight |  |  |  | 0,38* | 0,91* |

\*p-value < 1%;

The statistical analysis, Table 2, reveals the existence of significant associations, for p-value <1%, between the variables in the sample.

We should highlight the positive and significant correlations of the presence of intratumoral emboli with the presence of lymph node invasion and TNM staging. The formation of intratumoral emboli occurs due to tumor-induced coagulation changes. This event facilitates the spread of the disease, explaining its correlation with lymph node invasion [24].

Following the analysis of the results in Table 2, it shows that the TNM staging variable presents a positive and significant correlation with the lymph node invasion variable. This correlation was also expected, as the TNM staging calculation uses lymph node invasion as one of its parameters. These results show that the mathematical model used is in accordance with the clinical classification used to establish the TNM staging.

It is noted that the correlations involving the variables presence of intratumoral emboli, lymph node invasion and TNM staging, shown in Table 2, did not show significant connections with the variables age and BMI, so that the correlations presented describe risk factors independent of age and of the patients’ body weight at diagnosis. However, it is known that both age and obesity are considered determinant risk factors.

Table 2 shows a positive and strong correlation between menopause and the patient’s age. This is an expected association, as women experience hormonal failure with aging. On the other hand, the data do not show a significant correlation between menopause and overweight.

It is concluded that due to the heterogeneity of the behavior patterns of the clinical parameters evaluated in breast cancer, it is necessary to analyze the sample data in smaller groups. Thus, probably more specific correlations can be evidenced from the data.

A useful methodology for observing similar characteristics, which subgroups present within a group (full sample), can be implemented by calculating the distances between variables. In this context, the Euclidean distance is used to determine which data are more homogeneous among themselves, while the Ward’s hierarchical agglomerative method is a criterion used in hierarchical cluster analyses to find the most heterogeneous subgroups among them [17,19].

Considering the data from the complete sample, Ward’s method results in the formation of hierarchical groups by similarities with fusion level g = 347.806027, red line in Figure 2.

**Figure 2.**
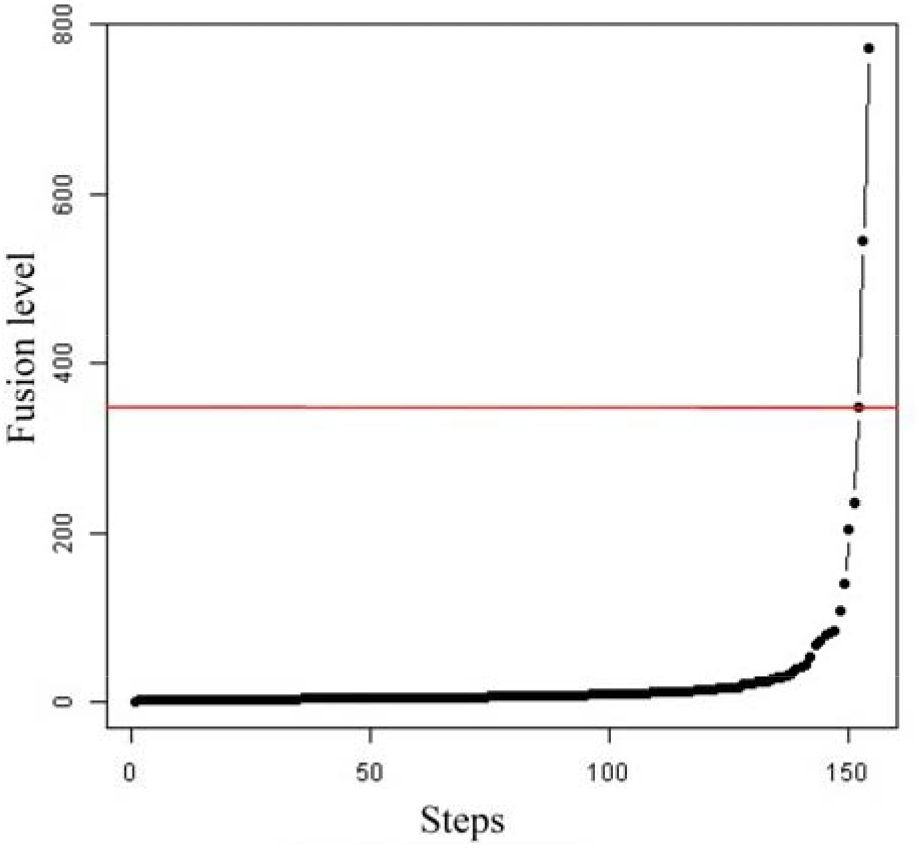
Fusion level at each step of Ward’s method.

The last three points on the curve in Figure 2 describe jumps in the algorithm’s steps, suggesting a marked reduction in similarity when 2 or 3 clusters are obtained. This result indicates that the algorithm needs to be completed in one of these steps.

To validate the results, scenarios with 2 and 3 clusters were studied, but little difference was observed between the groups. It was decided to keep 3 clusters, because clinically there was a better representation of information about patients.

The result of the analysis of Ward’s method, presented in the form of a dendrogram, Figure 3, helps to identify the division of clusters, denoted by C_1_, C_2_ and C_3_, respectively.

**Figure 3.**
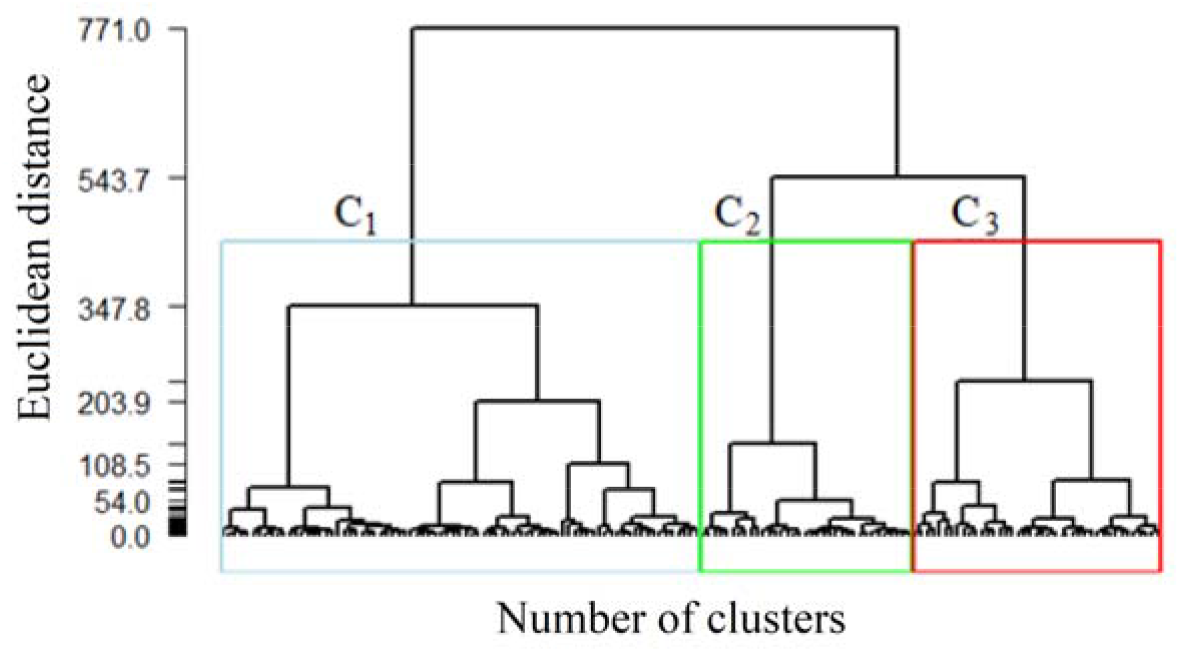
Hierarchical formation of groups by similarity, using Euclidean distances.

From the descriptive means and test of means, see Table 3, it appears that clusters C_1_, C_2_ and C_3_ do not show significant differences in the variables intratumoral emboli, presence of lymph node invasion, molecular subtype, grade, TNM staging, tumor size and height.

**Table 3.** Descriptive means and test of the means of variables for each cluster

| | Full sample | Cluster $C_1$ | Cluster $C_2$ | Cluster $C_3$ |
| --- | --- | --- | --- | --- |
| Number of patients | 155 | 79 | 35 | 41 |
| **Binary variables** |  |  |  |  |
| Presence of intratumoral emboli | 0,31 | 0,27<sup>a</sup> | 0,31<sup>a</sup> | 0,39<sup>a</sup> |
| Presence of lymph node invasion | 0,41 | 0,33<sup>a</sup> | 0,51<sup>a</sup> | 0,49<sup>a</sup> |
| Menopausal status | **0,70** | **0,99<sup>a</sup>** | **0,26<sup>c</sup>** | **0,51<sup>b</sup>** |
| **Categorical variables** |  |  |  |  |
| Molecular subtype | 2,43 | 2,34<sup>a</sup> | 2,66<sup>a</sup> | 2,39<sup>a</sup> |
| Grade | 1,93 | 1,95<sup>a</sup> | 1,91<sup>a</sup> | 1,90<sup>a</sup> |
| TNM staging | 2,06 | 1,96<sup>a</sup> | 2,09<sup>a</sup> | 2,22<sup>a</sup> |
| **Quantitative variables** |  |  |  |  |
| Tumor size (cm) | 3,18 | 3,11<sup>a</sup> | 3,51<sup>a</sup> | 3,03<sup>a</sup> |
| Age (years) | **56,64** | **67,06<sup>a</sup>** | **42,57<sup>c</sup>** | **48,56<sup>b</sup>** |
| Weight (kg) | **72,54** | **68,66<sup>b</sup>** | **62,59<sup>c</sup>** | **88,54<sup>a</sup>** |
| Height (m) | 1,60 | 1,59<sup>a</sup> | 1,61<sup>a</sup> | 1,62<sup>a</sup> |
| BMI (kg/m<sup>2</sup>) | **28,25** | **27,14<sup>b</sup>** | **24,11<sup>c</sup>** | **33,94<sup>a</sup>** |

Different letters indicate a significant difference between the means of a variable in the different clusters.

On the other hand, all clusters show significant differences in the menopausal status, age, weight and BMI variables. Thus, in our mathematical modeling, these variables were used to characterize their influence on breast cancer prognosis.

It is observed in Table 3 that cluster C_1_, composed of 79 patients, stands out for containing 99% of menopausal women, older, with a mean age of 67 years and mean BMI of 27.14 kg/m^2^, therefore considered overweight.

Cluster C_2_, represented by 35 patients, contains younger women with an average age of 42.57 years. Most are not in menopause, and 51% of them have lymph node invasion. The average BMI is 24.11 kg/m^2^considered a normal weight.

Cluster C_3_, composed of 41 patients, contains women of intermediate ages, with an average age of 48.56 years. These patients have a BMI of 33.94 kg/m^2^, that is, they are patients with grade 1 obesity. It is also observed that 51.21% of these women are in menopause and 49% have the presence of lymph node invasion.

Other information regarding the categorical and quantitative variables in each cluster are shown in Figures 4 and 5. The distributions in blue, black and red, represent the information about patients allocated in clusters C_1_, C_2_ and C_3_, respectively.

**Figure 4.**
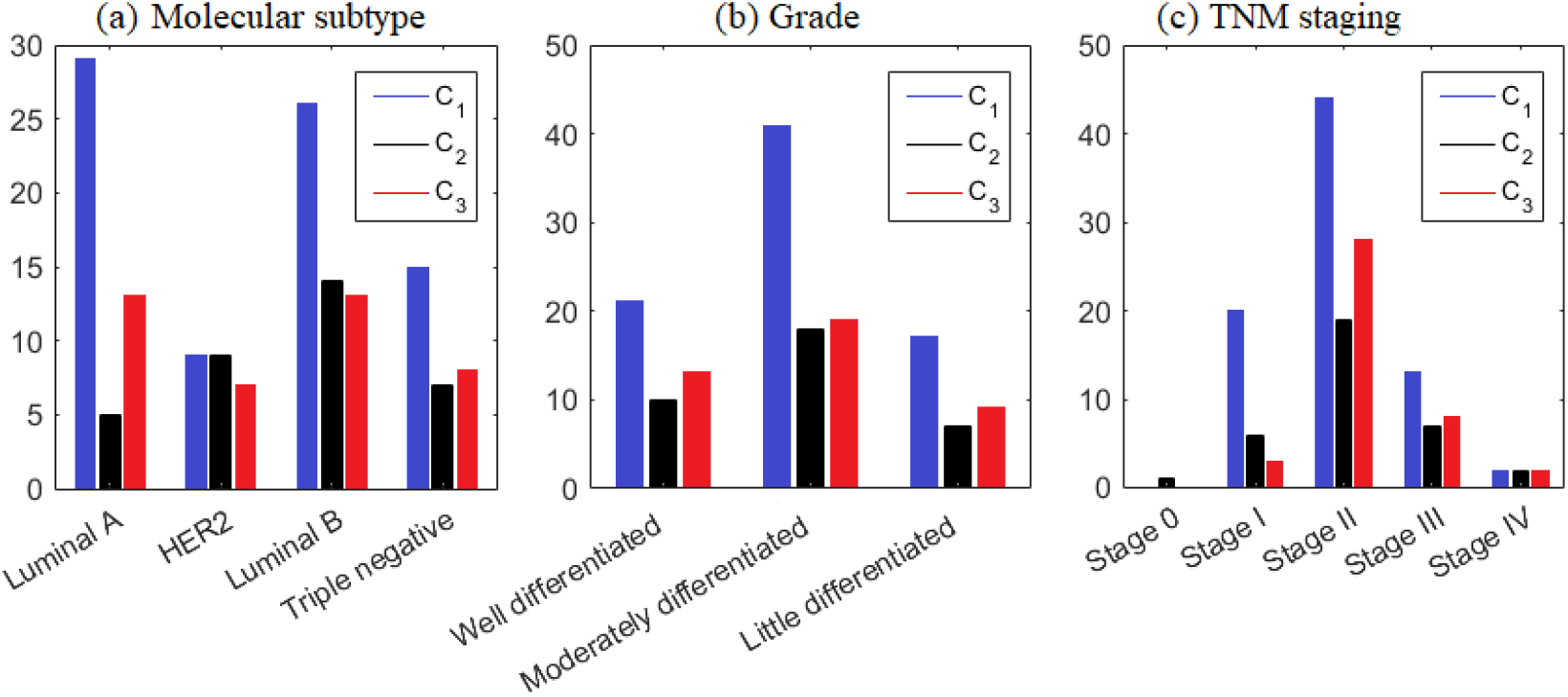
Distribution of categorical variables of clusters C_1_ (blue), C_2_ (black) and C_3_ (red).

**Figure 5.**
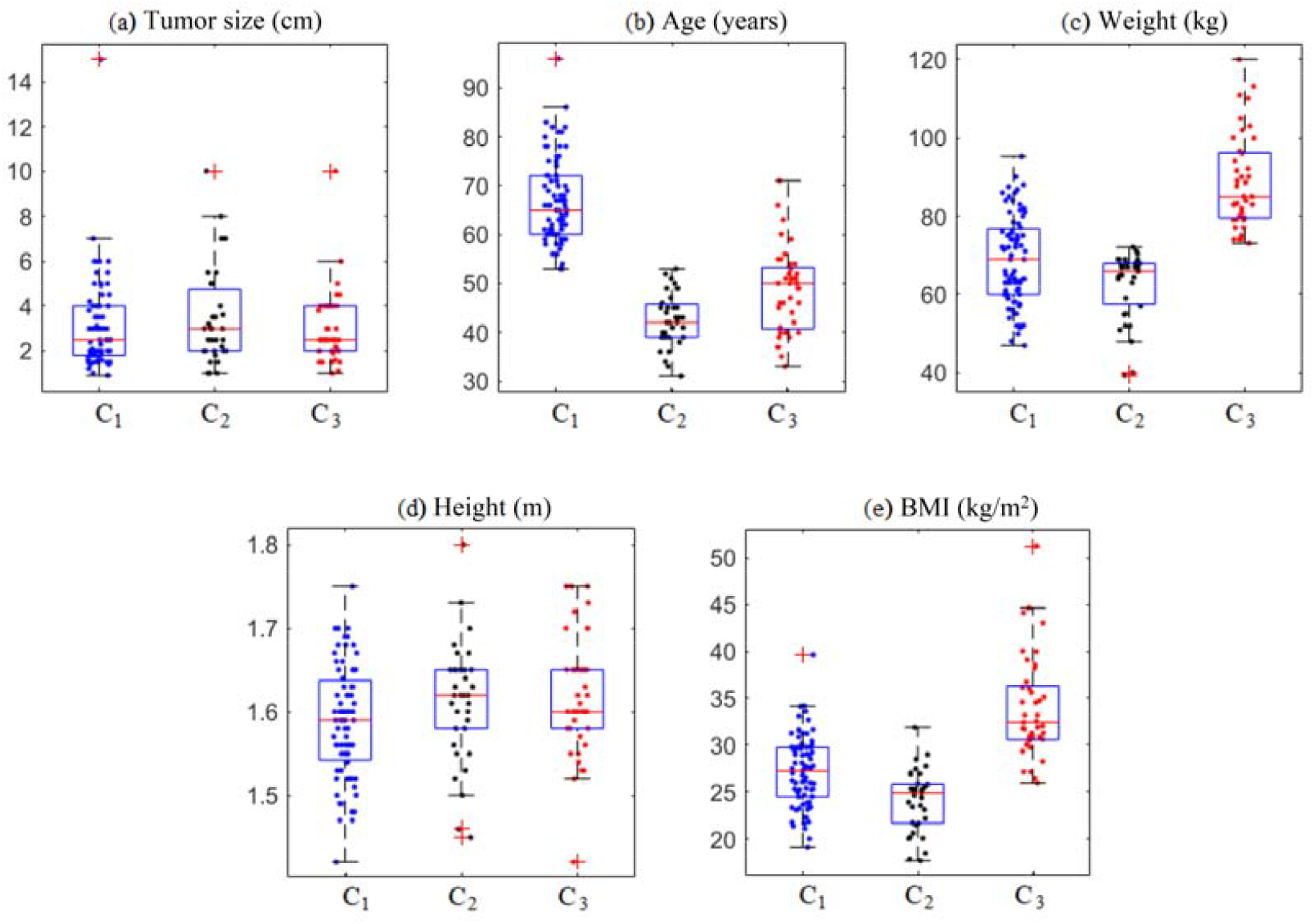
Distribution of quantitative variables in clusters C_1_ (blue), C_2_ (black) and C_3_ (red).

Figure 4 shows the prevalence of tumors of the Luminal A and B subtype in clusters C_1_ and C_3_, and Luminal B in cluster C_2_. This means that cluster C_2_ groups patients with the worst clinical prognosis when compared to the others, since Luminal B subtype tumors are quite aggressive [25].

Regarding the histological grade, most patients in the clusters have an intermediate grade, when the tumors are moderately differentiated. As for the TNM staging, most were in stage II for all groups.

Figure 5(a) shows that patients in clusters C_1_, C_2_ and C_3_ had tumors up to 7cm, 8cm and 6cm, respectively, with some outliers detected. Most older patients are found in cluster C_1_, as shown in Figure 5(b). The BMI of the patients ranged from 17.63 kg/m^2^ to 51.26 kg/m2, with 26.45% of the patients having a BMI below 25 kg/m^2^, 40.65% with a BMI between 25 kg/m^2^ and 30 kg/m2, and 32,9% with BMI above 30 kg/m^2^. Figure 5(e) shows that all patients in cluster C_3_ have a high BMI.

Thus, it is observed that with the division of clusters, through multivariate analysis, it was possible to characterize a heterogeneity of behavior between the clinicopathological variables. Thus, quantifying the intensity of the statistical dependence of the set of variables in each cluster will allow us to understand the influence that one variable exerts over another, making it possible to identify possible risk factors associated with the groups.

Therefore, using the calculation of Spearman’s linear correlation between the variables, the results presented in Table 4 are obtained.

**Table 4.**
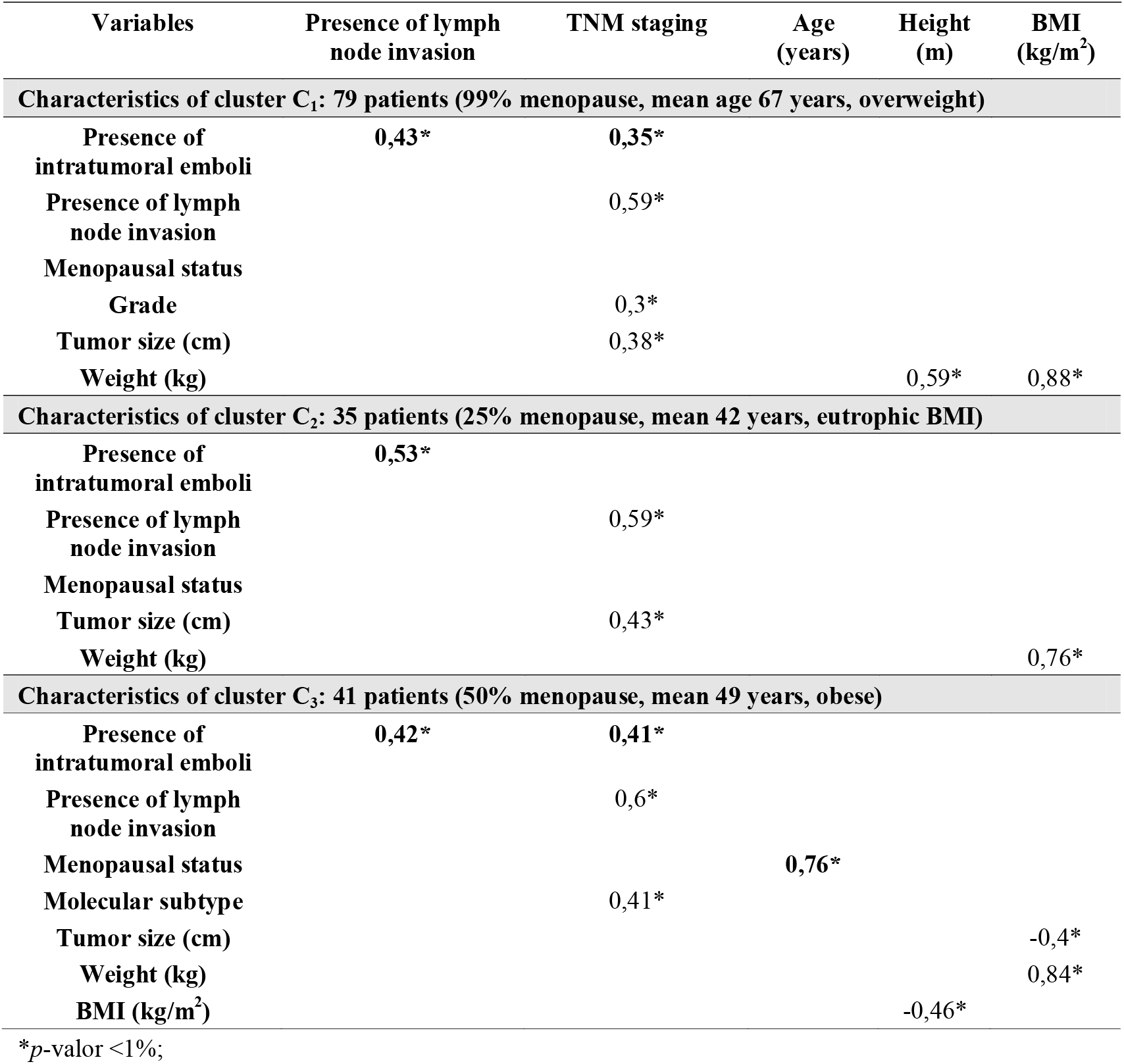
Spearman correlations of variables for clusters C_1_, C_2_ and C_3_.

| Variables | Presence of lymph node invasion | TNM staging | Age (years) | Height (m) | BMI (kg/m<sup>2</sup>) |
| --- | --- | --- | --- | --- | --- |
| **Characteristics of cluster C<sub>1</sub>: 79 patients (99% menopause, mean age 67 years, overweight)** |  |  |  |  |  |
| Presence of intratumoral emboli | 0,43* | 0,35* |  |  |  |
| Presence of lymph node invasion |  | 0,59* |  |  |  |
| Menopausal status |  |  |  |  |  |
| Grade |  | 0,3* |  |  |  |
| Tumor size (cm) |  | 0,38* |  |  |  |
| Weight (kg) |  |  |  | 0,59* | 0,88* |
| **Characteristics of cluster C<sub>2</sub>: 35 patients (25% menopause, mean 42 years, eutrophic BMI)** |  |  |  |  |  |
| Presence of intratumoral emboli | 0,53* |  |  |  |  |
| Presence of lymph node invasion |  | 0,59* |  |  |  |
| Menopausal status |  |  |  |  |  |
| Tumor size (cm) |  | 0,43* |  |  |  |
| Weight (kg) |  |  |  |  | 0,76* |
| **Characteristics of cluster C<sub>3</sub>: 41 patients (50% menopause, mean 49 years, obese)** |  |  |  |  |  |
| Presence of intratumoral emboli | 0,42* | 0,41* |  |  |  |
| Presence of lymph node invasion |  | 0,6* |  |  |  |
| Menopausal status |  |  | 0,76* |  |  |
| Molecular subtype |  | 0,41* |  |  |  |
| Tumor size (cm) |  |  |  |  | -0,4* |
| Weight (kg) |  |  |  |  | 0,84* |
| BMI (kg/m<sup>2</sup>) |  |  |  | -0,46* |  |

\*p-value <1%;

Table 4 confirms some expected characteristics, highlighting the strong correlation between the variables weight and BMI, in all clusters, and the correlation between the variables menopausal status and age at diagnosis, in cluster C_3_. Similarly, correlations between the presence of intratumoral emboli, presence of lymph node invasion and TNM staging are present in almost all clusters. It is worth mentioning that, despite these characteristics having already been observed in the data of the complete sample, see Table 2, it is now possible to analyze these correlations in the context of the particularities of each cluster. Next, the analysis of each cluster is carried out.

##### Cluster C_1_

In cluster C_1_, composed of menopausal, older and overweight patients, the presence of significant correlations between the variables intratumoral emboli, lymph node invasion and TNM staging are observed. This statement is justified by the analysis of the correlations obtained in Table 4, that is:

a. the correlation of intratumoral emboli with lymph node invasion and TNM staging,
b. the correlation of TNM staging with lymph node invasion, tumor grade and size.

It is known that, clinically, the formation of intratumoral emboli occurs due to coagulation alterations induced by tumors, facilitating the process of spreading the disease. Furthermore, the larger the size of the tumor, the more advanced the TNM stage of the disease is.

##### Cluster C_2_

Cluster C_2_ is composed of patients aged between 31 and 52 years, most of them not menopausal and with an average BMI of 24.11 kg/m^2^. There are significant correlations between the presence of intratumoral emboli and lymph node invasion, **without association with the obesity variable**. This statement is justified by the analysis of the correlations obtained in Table 4, that is:

a. the correlation of intratumoral emboli with lymph node invasion,
b. the correlation of TNM staging with lymph node invasion and tumor size.

Clinically, these correlations act in favor of the same biological event, which in this case would be favoring tumor spread. In addition, this association has an important clinical significance, since this cluster is characterized by the incidence of the disease in young women, which gives them a risk of occurrence of highly aggressive tumors [26]. Anyway, the fact that these women are not in menopause at diagnosis is another factor of worse prognosis, because estrogen acts as fuel for breast cancer [27].

##### Cluster C_3_

Cluster C_3_, composed of patients considered young, obese and with a prevalence of TNM staging in stages II and III, presents a strong correlation between the menopausal status variables and age at diagnosis, in addition to other correlations previously observed in the preceding clusters. This statement is justified by the analysis of the correlations obtained in Table 4, that is:

a. the correlation of intratumoral emboli with lymph node invasion and TNM staging,
b. the correlation of TNM staging with intratumoral emboli, lymph node invasion, and molecular subtype,
c. the correlation of menopausal status with age at diagnosis.

Clinically, the strong correlation between the variables menopausal status and age at diagnosis configures a worse prognosis of the disease for non-menopausal women. In these data, there is no correlation between obesity and variables associated with breast cancer. On the other hand, in the literature it is observed that obesity is a risk factor for the occurrence of breast cancer and is associated with the occurrence of highly aggressive tumors [28,29].

### Spatial distribution of clinicopathological variables

Table 5 presents the descriptive means of the variables considered, for each cluster, for each of the three regions of the 8th Health Regional of the State of Paraná, that is, for the regions of Fronteira, Vale do Iguaçu and Vale do Marrecas. The municipalities in these regions are shown in Figure 1.

**Table 5.** Means of the variables in each cluster for the regions of the 8th Health Regional of the State of Paraná.

|  | Fronteira |  |  | Vale do Iguaçu |  |  | Vale do Marrecas |  |  |
| --- | --- | --- | --- | --- | --- | --- | --- | --- | --- |
|  | C<sub>1</sub> | C<sub>2</sub> | C<sub>3</sub> | C<sub>1</sub> | C<sub>2</sub> | C<sub>3</sub> | C<sub>1</sub> | C<sub>2</sub> | C<sub>3</sub> |
| Number of patients | 30 | 10 | 15 | 17 | 5 | 9 | 32 | 20 | 17 |
| **Binary variables** |  |  |  |  |  |  |  |  |  |
| Presence of intratumoral emboli | 0,30 | 0,40 | **0,47** | 0,18 | **0,40** | 0,33 | 0,28 | 0,25 | **0,35** |
| Presence of lymph node invasion | 0,30 | **0,60** | 0,47 | 0,29 | **0,80** | 0,67 | 0,38 | 0,40 | 0,41 |
| Menopausal status | 0,97 | **0,10** | 0,33 | 1,00 | **0,40** | 0,67 | 1,00 | **0,30** | 0,59 |
| **Categorical variables** |  |  |  |  |  |  |  |  |  |
| Molecular subtype | 2,27 | 2,70 | 2,27 | 2,29 | 2,40 | **2,78** | 2,44 | 2,70 | 2,29 |
| Grade | 1,87 | **2,30** | 1,93 | 1,76 | 1,60 | 1,89 | 2,13 | 1,80 | 1,88 |
| TNM staging | 1,93 | 2,00 | 2,27 | 1,76 | **3,00** | 2,22 | 2,09 | 1,90 | 2,18 |
| **Quantitative variables** |  |  |  |  |  |  |  |  |  |
| Tumor size (cm) | 3,22 | 2,38 | 2,99 | 2,84 | **3,80** | 3,50 | 3,15 | **4,00** | 2,81 |
| Age (years) | 65,60 | 43,00 | 46,87 | 69,59 | **42,40** | 50,44 | 67,09 | **42,40** | 49,06 |
| Weight (kg) | 71,07 | 60,62 | 84,91 | 66,48 | 56,88 | 83,90 | 67,55 | 65,00 | **94,19** |
| Height (m) | 1,59 | 1,59 | 1,62 | 1,58 | 1,62 | 1,61 | 1,60 | 1,62 | 1,63 |
| BMI (kg/m<sup>2</sup>) | 28,24 | **23,92** | 32,76 | 26,63 | **21,58** | 32,58 | 26,37 | **24,84** | 35,70 |

Note that the means of the variables that stand out the most are those of cluster C_2_. Although the Vale do Iguaçu region contains the smallest number of patients, those in cluster C_2_ of this region showed that the youngest patients are in a very advanced stage of the disease, stage III, with the worst prognosis of the disease in all the sample. Here, new analyzes are suggested to assess the impacts of extrinsic factors to breast cancer patients, especially environmental factors, health habits and diet.

Table 6 shows the percentages of the spatial distribution of patients in the clusters. It is observed in Tables 5 and 6 that the lowest and highest frequency of patients with the disease occur in Vale do Iguaçu and Vale do Marrecas, respectively.

**Table 6.** Spatial distribution of the percentages of patients in clusters C_1_, C_2_ and C_3_ in the regions of the 8th Health Regional of the State of Paraná.

|  | Fronteira | Vale do Iguaçu | Vale do Marrecas |
| --- | --- | --- | --- |
| cluster $C_1$ | 37.97% | 21.52% | 40.5% |
| cluster $C_2$ | 28.57% | 14.28% | 51.1% |
| cluster $C_3$ | 36.58% | 21.95% | 41.46% |

Regarding the higher frequency of patients who developed the disease being in the Vale do Marrecas region, in the literature there are studies that associate this region with the highest values of pesticide use in the period from 2011 to 2016, in the state of Parana [30]. These studies highlights that this situation is considered serious in almost all municipalities in the 8th Health Regional of the State of Paraná [30]. Therefore, knowledge about the spatial distribution of pesticide use can be used as a variable for the prognosis of breast cancer, as well as to give a possible interpretation of the data presented in Tables 5 and 6.

## CONCLUSION

This study aimed to analyze data from patients with breast cancer, attended at a reference Hospital in Oncology (Francisco Beltrão Cancer Hospital) by the Unified Health System (SUS), in the period 2015-2020, in the southwest region of Paraná, Brazil, considering determinant clinicopathological variables for the prognosis of the disease.

It is observed in the data sample that 70% of the patients were in menopause at diagnosis, 31.5% had tumors containing emboli, and 41% had positive lymph nodes. The prevalence of Luminal subtype B tumors, intermediate histological grade, and TNM staging II was verified. Furthermore, the prevalence of the disease was higher in women aged over 50 years, representing 66% of cases. The BMI of the patients ranged from 17.63 kg/m^2^ to 51.26 kg/m^2^, with 26.45% of the patients with a BMI below 25 kg/m^2^, 40.65% with a BMI between 25 kg/m^2^ and 30 kg/m^2^ and 32.9% with BMI above 30 kg/m^2^.

Through statistical analysis it was possible to determine the heterogeneity of the data, so the patients were separated into three clusters. When analyzing the obtained clusters, each one of them had specific characteristics. Cluster C_1_ is formed by older, menopause and overweight women; cluster C_2_ by young, non-menopausal women with eutrophic BMI; while cluster C_3_ is formed by menopausal and obese women. Thus, with the division of the groups, it was possible to characterize a heterogeneity of characteristics of the clinicopathological variables. Quantifying the intensity of the statistical dependence of the set of variables in each cluster allowed us to understand the influence that one variable exerts over another, enabling the identification of possible risk factors associated with the groups.

Finally, an analysis of clusters using the spatial distribution of clinicopathological variables was presented. It was observed that the region of Vale do Iguaçu, despite containing the smallest number of patients, was the region with the worst averages for the clinicopathological variables, while the Vale do Marrecas region had the most cases of the disease.

## Data Availability

The sequential data used contain information from biopsy samples taken serially from women who had lesions suggestive of breast cancer, visualized by imaging tests (mammography, ultrasound, or MRI) and physical examinations, in the period from May 2015 to March 2020. Data confidentiality was maintained in accordance with clinical research guidelines.

